# Unrealistic optimism in the eye of the storm: Positive bias towards the consequences of COVID-19 during the second and third waves of the pandemic

**DOI:** 10.1101/2022.05.10.22274918

**Authors:** Ada Maksim, Sławomir Śpiewak, Natalia Lipp, Natalia Dużmańska-Misiarczyk, Grzegorz Gustaw, Krzysztof Rębilas, Paweł Strojny

**Affiliations:** Institute of Applied Psychology, Faculty of Management and Social Communication, Jagiellonian University in Kraków, ul. Prof. St. Łojasiewicza 4, 30-348 Kraków, Poland

**Keywords:** unrealistic optimism, positive bias, positive illusions, contracting COVID-19, severe consequences of COVID-19

## Abstract

Research conducted at the outset of the pandemic shows that people are vulnerable to unrealistic optimism (UO). However, the Weinstein model suggests that this tendency may not persist as the pandemic progresses. Our research aimed at verifying whether UO persists during the second (Study 1) and the third wave (Study 2) of the pandemic in Poland, whether it concerns the assessment of the chances of COVID-19 infection (Study 1 and Study 2), the chances of severe course of the disease and adverse vaccine reactions (Study 2). We show that UO towards contracting COVID-19 persists throughout the pandemic. However, in situations where we have little influence on the occurrence of the event, the participants do not show UO. The exceptions are those who have known personally someone who has died from a coronavirus infection. These results are discussed in terms of self-esteem protection and the psychological threat reduction mechanism.

---

It is common for people to make predictions about their future. While pessimists tend to contemplate the worst-case scenario, optimists believe that good things will happen to them (1). According to recent meta-analyses, optimism is believed to be associated with benefits of various types, including health and well-being (2,3). For example, optimism is associated with a lower risk of cardiovascular events and all-cause mortality (3). Some authors (3,4) suggest that future studies should be focused on evaluating the benefits of interventions that are aimed at reducing pessimism and promoting optimism. However, not all aspects of optimism are desirable or beneficial.

Dispositional optimism “is defined as the generalized positive expectancy that one will experience good outcomes” (5) and is mostly responsible for the above-mentioned benefits. Its dark side variant is so-called *unrealistic optimism*, a cognitive bias that makes people think that negative events are more likely to happen to others, and positive events are more likely to happen to them (6,7). Although some researchers (4) posit that unrealistic optimism functions as a positive illusion that helps people to cope with potentially threatening experiences by reducing anxiety, others (6,8–10) point to the maladaptive aspects of the optimistic bias. For example, unrealistic optimism may be related to developing some dire conditions such as coronary disease (5), alcoholism (9), breast cancer in women, and prostate cancer in men (6) as people have tendency to underestimate own risk of developing serious health problems. Unrealistic optimism is also correlated with risky and hazardous behaviours. People who perceive themselves as better drivers than others admit to violating speed limits (10), and young women who presume they are less likely than others to get pregnant are also less likely to use effective contraception methods—such behaviour could result in an unwanted pregnancy (8).

## Unrealistic optimism during the COVID-19 pandemic

Being unrealistically optimistic about one’s chances of being infected by coronavirus (and the ability to infect others) may lead to the illusion that obeying the strict policies imposed by the government is simply unnecessary in one’s case (11). As a result, unrealistic optimism could lead to reckless behaviours during the pandemic, such as ignoring the protective measures recommended by the World Health Organization (WHO; keeping a social distance, covering mouth and nose with a mask, avoiding crowded or indoor settings, etc.) which could lead to spreading the disease (1). The issue of unrealistic optimism has grown in importance in light of recent research on the perceived risk of infection during the COVID-19 pandemic. Dolinski and his colleagues (1) decided to verify if the imminent COVID-19 pandemic would stimulate the expression of unrealistic optimism. The researchers tested whether subjects would perceive that they are exposed to the disease to the same extent as the average person like themselves or if they would be affected by the unrealistic optimism (or the opposite – unrealistic pessimism) bias. The research of Dolinski and colleagues (1) was conducted in March 2020 when the media reported about the first people diagnosed with coronavirus in Poland. In their study, the pattern of unrealistic optimism in the face of the beginning of the COVID-19 pandemic emerged. Similar results were obtained in other European countries (France, Great Britain, Switzerland, and Italy) in February 2020 before the collapse of the healthcare system in Italy (12). People who were asked about their chances of getting infected were generally optimistic and assessed the personal risk of contracting coronavirus as lower than others.

Further research results also point to the implications of the positive bias for health-related behaviours (13,14). According to Oljača and colleagues (15), the optimistic bias may indeed influence attitudes towards compliance with restrictions. In a study conducted in Serbia, the participants who scored higher on the UOS–NLE subscale (measuring unrealistic optimism towards negative life events) assessed the risk connected with COVID-19 infection as lower and declared lower compliance with the pandemic restrictions. Similarly, Gordeeva and colleagues (16) found a positive link between defensive optimism (the tendency to diminish the risk of the emergence of negative events) and failure to comply with the stay-at-home rule in their study conducted in Russia in March and April 2020.

## Predictors of unrealistic optimism in the context of the COVID-19 pandemic

So far, the most elaborated theoretical model of unrealistic optimism has been formulated by Neil Weinstein (7). Below we refer to the five factors that, according to the Weinstein model (7), may have the most significant impact on unrealistic optimism during the pandemic. At least two factors should inhibit the tendency to the positive bias among people at pandemic risk: (a) *the perceived probability of the event* and (b) *the ease of recalling a stereotypical victim of a given situation*. The first one is inherently connected with the pandemic’s growth and increasing numbers of people contracting coronavirus. Higher perceived frequency (i.e., probability) in the general population should affect personal judgement of the risk but not necessarily others’ judgement. Thus, when the event is more frequent, the unrealistic bias may be weakened by a higher own risk rating. The second predictor that works in a similar direction means that the assumption regarding stereotype salience is based on the representativeness heuristic (17). Weinstein (7) assumes that the harder it is to imagine a typical victim of a specific event, the weaker the optimistic bias will be. As the pandemic spreads, individuals should be more aware that the severe consequences of COVID-19 affect not only the elderly with significant health problems but also younger, healthy people. With the increase in diversity and the number of victims of the pandemic, it will be more difficult to create a stereotypical image of the person most exposed to coronavirus, which should reduce the tendency to create cognitive illusions.

However, there are two other predictors that in our opinion would work in opposite directions to enhance positive illusions: (c) *controllability of the situation*, (d) *the degree of desirability*, and (e) *the personal experience* which the last one can work both ways. Controllability of the situation refers to a human’s sense that a situation’s outcome is dependent on their own actions. Therefore, people tend to overestimate their chances in positive events and underestimate their risks in negative events. In our opinion, people at risk during a pandemic may be vulnerable to the illusion of control through the availability of preventive measures: wearing a mask, keeping their distance, disinfecting hands. Thus, they can create the illusion of greater control of the situation and less chance of contracting the virus. In our opinion, a sense of control can foster a positive bias when asking individuals about the chances of contracting coronavirus. In this aspect, people seem more susceptible to the illusion of their own preventive actions but not necessarily for other aspects such as vulnerability to a severe course of COVID-19 or adverse vaccine reactions. The degree of desirability refers to the severity of the consequences. It is assumed that the more desirable a situation’s outcome, the greater the optimistic bias. However, negative events induce a more negative effect which leads to defensive strategies for protecting oneself and also results in higher optimistic bias. People desire positive outcomes, and when they are faced with the risk of losing their health or even their life they may be prone to reducing anxiety and protecting their self-esteem. One such strategy may be by creating positive illusions, which allows individuals to change their perception of a situation from threatening to less threatening. The last moderator mentioned by Weinstein (7) is the assumption about personal experience which is based on the availability heuristic (18). Previous experience with an event increases the belief in its reoccurrence. Personal experience may, in a similar vein, change the personal perception of the risk faced by an individual. However, two alternatives of these changes in the perception of the risk may be taken into account according to the existing literature (10).

The first one is the shift in the perception of the situation to be more threatening and even leads to the opposite unrealistic pessimism phenomenon (19) or to a lower level of unrealistic optimism (10). Alternatively, personal experience may, in some cases, result in enhanced self-protective motivation and may lead to an underestimation of the personal risk in relation to others (9).

## Aim of the studies

Considering the above, it seems important to examine the tendency to positive bias as the COVID-19 pandemic develops, so we decided to explore the susceptibility to unrealistic optimism during the second (Study 1) and the third wave of the pandemic in Poland (Study 2), when the number of infections increased dramatically. If the tendency to unrealistic optimism persists in the further stages of the pandemic, we expect to replicate the tendency to underestimate one’s own chance of contracting coronavirus despite the growth in the number of infections in the population both during the second (Study 1) and third (Study 2) waves of the pandemic in Poland. In addition, in Study 2, we decided to broaden the spectrum of assessing the tendency to unrealistic optimism with two issues that seem to be of particular importance as the pandemic develops: the severity of a potential COVID-19 infection and adverse vaccine reactions. As far as we know, there is scarce evidence whether the optimistic bias is limited only to the prediction of the chance of contracting COVID-19 or is related to other important health-related topics. The positive bias toward coronavirus risk assessments does not imply that people assume they are at risk of serious complications and at risk of losing their health and even their lives. On the contrary, it can be assumed that such cognitive bias may protect individuals from thinking about the serious consequences of contracting COVID-19. Thus, the presence of the cognitive bias towards contracting COVID-19 does not necessarily mean that people are positively biased towards assessment of the chances of a severe course of the disease or adverse vaccine reactions. Those two seem to be beyond individuals’ ability to take their own actions that could create the illusion of control and lead to the positive bias.

Based on the research by Dolinski et al. (1), to determine the sample size, we expected *Cohen’s d* effect sizes to be from *d* = 0.238 to *d* = 0.491. For the calculations, we adopted the average value of *d* = 0.365; expected power of .90 and alpha = .05. The analysis of the power in G*Power (version 3.1.9.7; 20) for the difference between the two dependent means and the two-tailed t-test, showed that the required power is achieved by a sample of 81 individuals. In order to meet these assumptions, we determined a sample size of at least 81 people in each of the studies.

We report all manipulations, measures, and exclusions in these studies (supplementary materials for more details). No studies in this manuscript were preregistered. All statistical procedures were performed in IBM SPSS v.26.0 (21)

## Study 1

We decided to conduct our first study in November 2020, during the spike of the very severe second wave of the pandemic in Poland. In 2020, 70,000 more people died in Poland than in previous years, which is nearly 20% more than in 2019 (and at the same time is the highest rate of death since World War II) (22). In November 2020 alone, 605,885 coronavirus cases were confirmed in Poland, and 11,494 people died as a result of the infection (23). Thus, the possible ramifications of excessive optimism became visible to every naive person. We expect to replicate the effect obtained by Dolinski and colleagues (1) who conducted their study at the beginning of the pandemic in Poland when people had not yet faced the widespread traumatic experiences of the deaths of their relatives and friends due to coronavirus.

## Method

This study was part of a larger research plan concerning decision-making about resource allocation during the coronavirus pandemic. In this report, we focus on the elements of the procedure related to the measurement of unrealistic optimism (a full description of the procedure and other measures used in the study can be found in the supplementary materials). Detailed information on the materials and instructions for Study 1 can be found in the repository: https://zenodo.org/record/5984642.

The study was approved by Research Ethics Committee at the Institute of Applied Psychology, Faculty of Management and Social Communication Jagiellonian University in Krakow. The participants were informed about the confidentiality of their data, the voluntary nature of the study and the possibility of ceasing to complete the survey at any time. Their answers were anonymized in the database. As the research was conducted online the consent to participate in the study was obtained online by entering personal data and clicking on the “continue” button.

### Participants

The first study involved 111 participants (90 female and 21 male) aged 18 to 42 years (*M* = 22.23, *SD* = 3.53). All of the participants were students of the Jagiellonian University in Kraków. Of all the participants, two were quarantined during the study, and four had been, at some point, diagnosed with COVID-19. The participants were assigned to the study conditions on the basis of quasi-randomization. At the very beginning of the study, they were asked about their day of birth. The numbers between 1 and 31 were divided into four intervals which were used to redirect the participants to two different conditions (control and experimental) based on their answers.

## Materials

### Pandemic and neutral context

The participants were assigned to one of two conditions related to one of two contexts—pandemic vs non-pandemic—as a part of the larger research project mentioned earlier. Four separate photos were used (i.e., two for the pandemic context and two for non-pandemic). More information about the chosen photos can be found in the supplementary materials. We had no theoretical predictions about the impact of manipulating the context (i.e., pandemic vs. neutral), however, due to the fact that context can significantly change the perception of the social situation, especially in people who are more or less exposed to the effects of a pandemic, we decided to take it into account in our preliminary analyses.

### Unrealistic optimism measurement

Our main dependent variable was the measure of how the participants were unrealistically optimistic about the possibility of contracting coronavirus. To this end, we asked them two questions:

1. How would you rate your chances of contracting coronavirus?
2. How would you rate the chances that someone else like you will contract coronavirus?

The participants answered both questions by estimating the chance of becoming infected as a percentage (from 0 to 100). It is suggested by Harris et al. (24) that such an indirect method of assessing the optimistic bias (by asking two separate questions) is more beneficial and informative than the classical, direct way introduced by Weinstein (7) which consists of only one item where participants are asked to compare themselves to an average other.

### Procedure

The first study was conducted in November 2020 at the peak of the second wave of the coronavirus in Poland. Due to pandemic restrictions, the study was conducted online. Participants received an email invitation to take part in the study. If they clicked the link received in the email, they were redirected to an online survey. Firstly, they were informed about data privacy and gave active, informed consent. Then, after assignment to research conditions, they were asked two questions regarding the perceived chance of contracting coronavirus—by themselves and someone similar to them. Finally, the participants filled in demographic data and were thanked for participating in the study. Detailed information about all the additional materials and scales used in the study can be found in the supplementary materials.

## Results

Due to the fact that being infected with COVID-19 at some point could influence the assessment of the risk of contracting coronavirus, participants who had been diagnosed with COVID-19 (*n* = 4) and participants who were in quarantine (*n* = 2) were excluded from the analysis. Preliminary analysis (general linear model with the assessment of the chance of infection as a within-subject factor and sex, age, and condition as between-subject factors) showed that the assessment of the chance of infection was not influenced by the experimental condition (see pandemic vs neutral photos; *F*(1, 79) = 0.07, *p* = 0.791. There were also no differences related to the age (*F*(12, 107) = 1.53, *p* = 0.130) and sex (*F*(1, 79) = 0.03, *p* = 0.869) of the participants. Therefore, in further analysis, all results were considered jointly, regardless of the manipulation of the pandemic vs the non-pandemic context, age, and gender of the participants. The estimate of contracting coronavirus oneself was significantly different from the estimate of it being contracted by someone else. The participants assessed their chance of becoming infected (*M* = 52.97, *SD =* 24.24) as lower than the chance of someone else becoming infected (*M* = 61.18, *SD* = 23.26). This difference is statistically significant and effect size is of moderate strength (*t* (103) = -4.69; p <0.001, *Cohen’s d* = -0.34).

## Discussion

The results of our first study correspond with other reports confirming a tendency to unrealistic optimism in the context of the assessment of life-or health-threatening events (5,6,9). It can be argued that a similar relationship may occur in people’s behaviour in response to the threat of the COVID-19 pandemic. Our study is in line with the few reports to date (1,12– 15), that show that unrealistic optimism may bias those who are at risk of the coronavirus pandemic. For example, in the study by Dolinski and colleagues (1), which was carried out at the beginning of the pandemic in Poland, the same group of students was asked three times about their assessment of the chances of contracting coronavirus, and it was shown that the tendency to unrealistic optimism remained stable among men but actually intensified among women during the week after the first COVID-19 infection was announced. However, it is not clear—according to the Weinstein model (7) — whether the tendency to underestimate the chances of contracting coronavirus will continue over the long term. Our study initially confirmed that there was a continuing tendency to underestimate the chances of catching COVID-19 during an exacerbating pandemic in Poland.

### Study 2

In our second study, we decided to extend the scope of the optimistic bias exploration to more specific aspects of pandemic risk. During the development of a pandemic, two aspects seem to be particularly important, and little known from the point of view of unrealistic optimism: (a) estimating the chances of serious complications as a consequence of a possible COVID-19 infection, and (b) the perceived risk of developing adverse vaccine reactions. There is strong evidence in the literature on unrealistic optimism suggesting that this effect occurs rather in the case of events that we assume we can control to some extent (1,7). According to Weinstein (7), in the case of events that people feel they can control it is easier for them to visualise their own behaviour aimed at reducing the risk. Thus, they overestimate their influence on the situation and are more susceptible to the optimistic bias. People may, to some extent, try to minimise the risk of contracting COVID-19 through their behaviour, thus, the possibility of becoming infected seems to be dependent on a person’s actions and under their control. However, people believe that they have no control over whether, as a result of the infection, they will experience serious complications that may result in death or a serious threat to life. Thus, we hypothesise that although people will underestimate the chances of getting ill, at the same time they will not underestimate the chances of developing a serious course of the disease as a consequence of a possible COVID-19 infection.

The chances of getting infected can be effectively reduced by following the recommendations of the WHO: limiting social contacts, wearing a face mask, or disinfecting hands. In the event of a severe course of COVID-19 infection, people do not have personal control over how the disease develops. In a study by Asimakopoulou and colleagues (11), participants showed lower unrealistic optimism when asked about the risk of hospitalisation, being taken into the intensive care unit, and being ventilated due to COVID-19 (less manageable situations) than when asked about the risk of contracting coronavirus or infecting someone else (more manageable situations). Therefore, we assume that in the case of the risk of a severe course of COVID-19, the effect of unrealistic optimism will be weaker.

We decided to include one more variable which was not included in previous studies as they were conducted during a different stage of the pandemic. Our second study was conducted in February 2021, after the second coronavirus wave in Poland, which turned out to be much more severe than the first one. Between September and December 2020, 1,205,878 new cases of coronavirus infections were confirmed and 25,656 people died due to a COVID-19 infection. At the peak of the second wave, COVID-19 patients occupied 20,000 hospital beds (23).

We assumed that, at this point, most of the participants will already have had their own experiences related to coronavirus, and in particular, they might personally know someone for whom contracting coronavirus had serious consequences. We decided to verify if the personal experience of knowing someone who had developed a severe illness due to COVID-19 or died from it would affect the unrealistic optimism of the participants.

In the literature on unrealistic optimism, we found mixed results related to the influence of personal experience (9,10). In a study by McKenna and Albery (10), participants who were involved in a car accident showed lower unrealistic optimism concerning their driving skills than other participants, but only if they were hospitalised as a result of the accident. In contrast, in a longitudinal study related to alcohol abuse (9), people who experienced negative consequences related to alcohol consumption at subsequent stages of the study still rated their own risk of developing serious problems related to alcohol abuse as lower than others.

Finally, since the date of the study coincided with the commencement of the vaccination programme in Poland, we were also interested in the assessment of the chances of adverse vaccination reactions – self versus others. We assumed that as the chances of adverse vaccination reactions are beyond one’s control, we will not observe optimistic bias in this case.

## Method

### Participants

The second study involved 84 participants (57 female, 26 male, and 1 nonbinary), aged from 19 to 65 (*M* = 35.42, *SD* = 9.07). Eleven of the participants were at some point diagnosed with COVID-19, two were quarantined while participating in the study. Out of 84 participants, 78 knew someone diagnosed with COVID-19, 47 knew someone who manifested severe symptoms of COVID-19, and 30 participants knew someone who died due to COVID-19.

### Materials

#### Pandemic and neutral context

Similarly, as in Study 1, we used the manipulation of the context of the unrealistic optimism assessment. Before the assessment, half of the participants were presented with pandemic-associated, death-related pictures whereas the other half were presented with - the same as in Study 1 - neutral images. The materials were chosen based on the separate pilot study. The stimuli used in the second study can be found in the supplementary materials. Detailed information on the materials and instructions for study 2 can be found in the repository: https://zenodo.org/record/5984642.

#### Unrealistic optimism measurement

As we wanted to verify whether unrealistic optimism would also apply to other aspects related to the coronavirus pandemic (apart from the assessment of the chances of being infected), we additionally asked the participants the following questions:

1. How would you rate your chances of a severe course of the disease if you contract coronavirus?
2. How would you rate the chances of someone else becoming severely ill if they contract coronavirus?

These questions related to the possible unrealistic optimism about a severe course of coronavirus disease. As the coronavirus vaccination programme was already underway during the second study, we also wanted to check if there were some differences in the assessment of the possible side effects of a vaccination:

1. How would you rate your chances of developing severe side effects after a coronavirus vaccination?
2. How would you rate the chances of someone else developing severe side effects after a coronavirus vaccination?

#### Personal experience of COVID-19

The second study was conducted on the verge of the third wave of the coronavirus pandemic in Poland. Thus, we assumed that the participants may have had some personal experience of COVID-19 at this point, which might have influenced the way they assessed their chances of getting infected and developing severe symptoms of COVID-19. At the end of the study, participants reported if they personally knew someone diagnosed with COVID-19, if they personally knew someone who developed severe symptoms of COVID-19, and if they personally knew someone who died due to a COVID-19 infection.

### Procedure

Again, due to the pandemic restrictions, the study was conducted online. Participants received an email invitation to take part in the study. If they clicked on the link received in the email, they were redirected to the online survey. Firstly, they were informed about data privacy and gave active, informed consent. Similar to the first study, participants were assigned to the research condition quasi-randomly, based on their day of birth. Depending on the condition, participants saw either neutral photos (control conditions) or photos related to the coronavirus pandemic (experimental conditions).

In the next step, they were asked to estimate their perceived chances of being infected with COVID-19, developing severe symptoms of COVID-19, and suffering severe side effects of vaccination against COVID-19. To assess the tendency to the optimistic bias, they also answered the same questions regarding their co-workers/other students. Finally, the participants filled in demographic data and information about their personal experience of COVID-19. More detailed information about the other measures used in the study can be found in the supplementary materials.

## Results

As in the first study, we excluded participants who declared that they had tested positive for the presence of coronavirus (*n* = 11) and participants who were in quarantine (*n* = 2) as their answers may have biased the results. Since during the second study, vaccination against coronavirus was already underway in Poland, we also excluded participants who had been vaccinated with at least one dose (*n* = 2). First, in the preliminary analysis, we checked if there were any differences in unrealistic optimism measures due to different experimental conditions (pandemic vs neutral photos). As we found none (*F*(1,15) = 0.13, *p =* .726), we decided to analyse all the data together. There were also no differences concerning unrealistic optimism due to the gender (*F*(1,15) = 0.13, *p =* .865) and age (*F*(32,15) = 0.71, *p =* .795) of the participants. The effect of unrealistic optimism related to the chances of contracting coronavirus has been successfully replicated (*t*(70) = -5.69, p < .001, *Cohen’s d* = -0.37). The respondents assessed their chances of becoming infected lower (*M* = 42.55, *SD* = 24.67) than the chances of other people (*M* = 51.03, *SD* = 22.98).

There was no effect of unrealistic optimism related to a severe course of COVID-19 infection (see Table 1). However, when assessing the chances of a severe course of the disease, personal experience related to coronavirus turned out to be an important factor. There was an interaction effect between unrealistic optimism and personal acquaintance with someone who died from a COVID-19 infection (*F*(1,68) = 6.50, *p* = .013, *Cohen’s d* = 0.58). Participants who knew someone who died as a result of COVID-19 infection estimated their chances of a severe course of coronavirus infection significantly lower (*M* = 26.29, *SD* = 19.74) than the chances of substantial side effects of COVID-19 infection for other people (*M* = 36.41, *SD* = 19.27; *p* = .028). This effect did not appear in the case of participants who did not personally know any victims of COVID-19 infection (*M*_*self*_ *=* 33.65 *SD*_*self*_ *=* 26.47 comparing to *M*_*other*_ *=* 29.61 *SD*_*other*_ *=* 22.47; *p* = .218).

**Table 1.** Summary of self and others’ chances assessments from the two studies

| How do you assess the chances<br>of... related to Covid-19 |  | Self |  | Others |  | Paired-samples *t*-test |  |  |  |  |
| --- | --- | --- | --- | --- | --- | --- | --- | --- | --- | --- |
|  |  | *M* | *SD* | *M* | *SD* | *t* | *df* | *p* | 95% *CI* |  |
|  |  |  |  |  |  |  |  |  | *Lower* | *Upper* |
| Study 1 | Contracting | 52.97 | 24.24 | 61.18 | 23.26 | 4.69 | 103 | <0.001 | -11.68 | -4.74 |
| Study 2 | Contracting | 42.55 | 24.67 | 51.03 | 22.98 | 5.69 | 70 | <0.001 | -11.45 | -5.51 |
|  | Serious complications | 31.13 | 24.48 | 31.94 | 21.53 | 0.297 | 69 | 0.768 | -6.28 | 4.65 |
|  | Developing adverse vaccine reactions | 27.06 | 24.43 | 28.33 | 22.14 | 0.912 | 69 | 0.365 | -4.05 | 1.51 |

Other experiences with the coronavirus pandemic (i.e., knowing people who have become infected or who have been severely ill) did not contribute to the effect of unrealistic optimism concerning a severe course of COVID-19 disease. There was no interactional effect in the case of a personal acquaintance with someone who has been severely ill (*F*(1,68) = 2.03, *p =* .158, *Cohen’s d* = 0.31). Additionally, we decided not to perform analysis on a personal acquaintance with someone who has been infected as a between-subject factor since 78 of 84 participants knew someone who has been ill.

There was also no effect of unrealistic optimism concerning potential adverse reactions of the COVID-19 vaccination (see Table 1). Overall, respondents rated the chances of experiencing vaccination side effects as low for themselves (*M* = 27.06; *SD* = 24.43) as for others (*M* = 28.33, *SD* = 22.14). Any type of personal experiences with the coronavirus pandemic were of no importance in this case (knowing someone who has been severely ill: *F*(1,68) = 0.32, *p =* .573; *Cohen’s d* = 0.14; knowing someone who died from COVID-19 infection: *F*(1,68) = 1.97, *p =* .166; *Cohen’s d* = 0.33).

## Discussion

The evidence presented in our research supports the assumption that the optimistic bias is maintained as the COVID-19 pandemic progresses and is not limited to the initial stages of a pandemic outbreak. Bottemanne and colleagues (25) suggest that the optimism bias may diminish as coronavirus spreads around the world. They argue that in face of an inevitable threat people tend to use unfavourable information more likely to update their beliefs. At first, the coronavirus pandemic was rather distant and novel but with more and more cases the risk of infection was getting higher and, as a result, this might have updated people’s beliefs about their personal chances of getting ill and weakened the optimistic bias. In contrast to the assumptions of Bottemanne and colleagues (25), our data, collected in two studies conducted during the second and third waves of the pandemic in Poland, confirm the existence of unrealistic optimism regarding the assessment of the chances of contracting coronavirus. Moreover, as the pandemic progressed, not only did the optimism not diminish, but the strength of the effect appears to be stable (i.e., *Cohen’s d* in Study 1 vs. Study 2 is 0.34 and 0.37, respectively). In both studies, the pandemic and non-pandemic contexts did not affect the assessment of any aspects of pandemic risk. This may be an argument for the high availability of information about the pandemic and relative insensitivity to additional information that would change the perception of reality during the second and third waves of the pandemic in Poland.

Interestingly, in the second study, people assessed both their own likelihood of becoming infected and of others as lower than in the first study (i.e., 52.97 vs 42.10 for one’s own assessment and 61.18 vs 51.00 for others). However, it is difficult to draw conclusions about the differences in the estimates of absolute values on that basis, because the results come from different groups of respondents at different stages of the pandemic’s development. We do not know whether this result indicates the opposite trend to that observed in the studies by Dolinski et al. (1) or whether it represents differences in the perceived probability of infection of various groups of people.

Nevertheless, maintaining the illusion of the lower vulnerability to infection that accompanies the sharp increase in the number of infected people — as we are dealing with in the second and third waves of the pandemic — indicates a strong cognitive bias that does not seem to have been reduced by the incoming information. According to Weinstein’s assumption of *the perceived probability of the event* (7), in March 2020, in Poland, there were a dozen new COVID-19 cases a day, while in the second half of the year, the numbers were oscillating around several thousand cases a day and more. As the incidence of the disease increases with the duration of the pandemic, individuals are supposed to make more realistic estimations of the chances of their own illness and should make those assessments similar to others, thus one would expect that the tendency to unrealistic optimism should decrease. However, the above argumentation assumes that people rationally evaluate the chances of positive and negative events in their lives, which, as we know from the many studies in the field about decision making and judgement, is no longer true (18,26). Likewise, the assumption that a growing number of infections should change the stereotypical image of a typical victim (7), which in turn should inhibit the tendency to the positive bias also turned out not to be valid in our studies. While people in our research showed positive illusions about coronavirus infection, the attempt to explain this phenomenon should focus on the role of factors that, from the theoretical point of view, could contribute to their maintenance.

One reason why individuals may be motivated to maintain a positive illusion is when they are trying to control an unpredictable situation (27). The outbreak of a pandemic is undoubtedly a factor that increases the unpredictability and uncertainty of actions and raises many risks related to the consequences of the decisions that individuals are making. There is a growing body of literature suggesting that the experience of uncontrollability increases uncertainty (28) and leads to the experience of lack of control which is challenging for various aspects of human functioning (29). In the context of our research, the most interesting seems the self-protection motivation (30,31) and regaining control when people are facing unpredictable situations (32). The positive illusion may be a form of self-protection and cognitive bias can serve as promoting positivity in one’s self-views. Following this argument, it can be expected that the growing number of infections will not only reduce the positive bias but will foster uncertainty about the future and enhance motivation to regain control of the situation, especially in terms of those aspects that may be perceived as controllable. We expected, according to Weinstein’s model (7), that the positive illusion will be especially strong in the case of the relatively controllable aspect of the pandemic situation (the chances of contracting of COVID-19) but not for those aspects that are beyond control (a severe course of COVID-19, adverse vaccine reactions). The results of Study 2 are consistent with the above assumptions and other studies suggesting the existence of the positive illusion for manageable rather than unmanageable situations (11). We predicted that in the case of the chances of infection, such an illusion of control is more likely to occur than for other aspects of assessment. Hand disinfection, self-isolation, and wearing a mask are actions that an individual can take at any time because they depend solely on their will. There is, however, an interesting contradiction in this aspect. Paradoxically, research shows that unrealistically optimistic people less often follow the rules and respect limitations. In fact, they tend to ignore protective measures and thus contribute to the spread of the virus. A positive illusion can therefore be a knife that cuts both sides: from an individual’s perspective, the belief that preventive measures are readily available strengthens the illusion of control, but it actually leads to the ignoring of limitations, which not only does not reduce the risk but also seriously increases it.

We did not expect to report the unrealistic optimism in regard to a severe course of COVID-19. The degree of desirability which refers to the severity of the consequences according to the Weinstein model (7) increases the pressure for a more realistic risk assessment. As the risk of a wrong and inadequate assessment of the situation increases, individuals pay higher costs for their wrong decisions. The results obtained are consistent with our assumptions that an individual will not be prone to unrealistic optimism when assessing a serious course of the disease. However, there is an interesting exception regarding people who knew someone who died from a COVID-19 infection. The results obtained in Study 2 show that people who experienced the death of a person they knew are unrealistically optimistic in regards to the assessment their own chances of a severe course of COVID-19. Knowing a person who died of COVID-19 may indicate the role of personal experience in the development of the positive illusion. The existing literature does not allow for conclusive assumptions about the influence of personal experience in the development of the positive bias. Rather, our results may suggest that personal experience enhances the positive illusion, however, there is another important factor that one cannot ignore. There is a considerable number of empirical findings suggesting the consequences of mortality salience evoke a psychological defence mechanism to protect self-esteem and reduce the psychological threat and anxiety (33,34). The personal experience in our study took a specific form beyond knowing someone infected with COVID-19. During the third wave of the pandemic, almost all respondents knew someone who had already been infected with coronavirus and our results suggest that those kinds of experiences are not sufficient to enhance the positive bias towards a severe course of the disease. The experience of COVID-19 does not necessarily imply its seriousness. In the case of death, we are dealing not only with the experience of severe complications but, above all, with mortality salience which bears far more psychological consequences (35) than only the experience of a severe course of coronavirus infection. Unfortunately, our study does not allow us to make a conclusion about the role of the specificity of these kinds of personal experiences. More research is needed to verify the role of assessing the consequences of infecting others in creating a positive illusion about the seriousness of the disease. It cannot be ruled out that unrealistic optimism may be a specific consequence of the awareness of one’s own mortality, which has not been verified in the empirical research so far.

In our research, we refer to the predictions based on the Weinstein model (7), which we consider to be the most elaborated theoretical framework in the literature explaining the predictors of unrealistic optimism. We are aware that the inference about the relationship to risk assessment in our research was indirect rather than direct. Further efforts should be made to better demonstrate the direct relationship of factors in the Weinstein model (7) with the development of the positive illusions regarding the assessment of various aspects of pandemic risk (contagion risk, risk of a severe course, risk of unexpected vaccine reactions) and the role of mortality salience in upholding positive illusions.

## Data Availability

The data underlying the results presented in the study are available in repository: https://zenodo.org/record/5984642.

